# Barriers and facilitators to implementing core osteoarthritis treatments in China: a mixed-method study

**DOI:** 10.1101/2025.01.10.25320307

**Authors:** Ziru Wang, Shuning Duan, Xier Chen, Huili Deng, Yunqi Wang, Guoxin Ni

**Affiliations:** PhD candidate, School of Sport Medicine and Rehabilitation, Beijing Sport University, Beijing, China; Master’s candidate, School of Sports Medicine, Wuhan Sports University, Wuhan, China; Master’s candidate, School of Medicine, Xiamen University, Xiamen, China; Masters, School of Physical Education and Health, Longyan University, Longyan, China; PhD, Department of Rehabilitation Medicine, the First Affiliated Hospital of Xiamen University, School of Medicine, Xiamen University, Xiamen, China

**Keywords:** Osteoarthritis, barriers and facilitators, implementation science, exercise therapy, qualitative, survey

## Abstract

**Objective:** To understand current practices and identify barriers and facilitators to implementing guideline recommended core OA treatments in China.

**Methods:** An exploratory mixed-methods design was employed, involving healthcare professionals managing OA in mainland China. The qualitative phase included semi-structured interviews (n=15) and a qualitative survey (n=181). A quantitative survey (n=302) evaluated the clinical applicability of identified themes, common practices, knowledge, and self-rated confidence in core capabilities about OA. Qualitative data were analyzed through thematic and content analysis using NVivo, and descriptive statistics were applied to quantitative data using RStudio.

**Results:** Participants representing six professions provided their perspectives qualitatively. Five themes emerged as barriers: widespread misconceptions about OA; limitations in current medical insurance policies; insufficient multidisciplinary collaboration; lack of workplace support; and low patient adherence to self-management. Three themes emerged as facilitators: telehealth and community-based delivery pathways; professional training and patient education resources; and personalized services with positive feedback.

Across 19 provinces in China, participants representing seven different health professions completed the quantitative survey. The most commonly used treatments for OA were physical agent therapy (56%), while exercise therapy was utilized by only 9%. The average OA knowledge score was 31.2 (±8.9) out of 55. Self-rated confidence was lowest for “referrals and interdisciplinary collaboration” (3.4±0.1, “somewhat confident”). The most applicable factors impacting the implementation of core OA treatments included patient comorbidities, knowledge of pain science and exercise therapy, and financial support (all 2.8±0.8, “applicable”).

**Conclusion:** Core OA treatments recommended by international clinical guidelines are infrequently implemented in China.

**SIGNIFICANCE AND INNOVATIONS:**

- This study is the first to explore the barriers and facilitators to implementing core osteoarthritis (OA) treatments in China, providing comprehensive insights into the unique challenges within the Chinese healthcare system.
- It highlights the substantial underutilization of exercise therapy in OA care (9%), reflecting a potential mismatch between guideline recommendations and clinical practice, and underscores the need for targeted implementation strategies.
- Major barriers include widespread misconceptions about OA, limited multidisciplinary collaboration, and insufficient financial and organizational support for non-pharmacological interventions.
- The study emphasizes the potential of telehealth and community-based care to address disparities in OA management and improve access to evidence-based, guideline-recommended OA treatments in diverse clinical settings across China.

---

Osteoarthritis (OA) is a multifactorial joint disease that is highly prevalent worldwide, affecting 133 million people in China as of 2019 ^1^. It imposes a burden on the individual, such as pain and physical dysfunction, as well as socio-economic burden, including healthcare costs and workforce loss ^2, 3^. By 2044, the OA burden in China is projected to increase 1.5 times from 2019, mainly driven by an aging population, rising obesity rates, and the country’s large population size ^4, 5^. Currently, there is no cure for OA ^6^. Addressing this major public health challenge in China is therefore crucial, and treatment priorities should focus on addressing OA symptoms rather than pathophysiological joint changes ^7^. High-quality clinical guidelines from the US, Australia, the UK, Europe, and international research societies consistently recommend exercise, education, and weight loss (where appropriate) as core treatments ^8–13^ for all people with OA, regardless of age, pain severity, or disease progression. However, it is widely acknowledged that people living in high-income countries receive suboptimal OA care ^14–16^, and implementing evidence-based OA care in low- and middle-income countries presents even greater challenges ^17^.

There has been no research evaluating the clinical practice patterns of healthcare professionals managing OA in China. This is important, given the recommendations of Chinese OA clinical guidelines and the structure of the Chinese healthcare system. In contrast to international OA guidelines which advocate exercise, education and weight loss (for overweight/obesity) in all people with OA, the 2024 edition of the Chinese OA clinical guideline^18^ adopts a stage-based treatment approach, recommending these core treatments only for patients in the Unlike in many Western countries, where accessing specialists often requires a referral from a primary care provider, China’s healthcare system allows patients to directly choose their first point of care. Patients can register to see any specialists in any hospitals without a referral, or choose to see generalist doctors at Community Health Centers (CHCs) for primary care ^19^. Despite the existence of allied health systems in other countries where physiotherapy is well-established and integrated into healthcare, the role of physiotherapists in China remains unstructured. For example, the term “physiotherapist” is often used interchangeably with “rehabilitation therapist,” creating confusion about their specific responsibilities and qualifications. Rehabilitation medicine is an emerging discipline in China, with its formal application beginning only in 1982, and it remains in the developmental stage within China’s medical service system ^20, 21^.

Successful implementation of high-quality evidence into clinical practice necessitates a comprehensive understanding of current practice patterns, along with barriers and facilitators experienced by the stakeholders in specific settings, in order to identify strategies to improve implementation ^22^. While research in other countries has already advanced to focus on OA models of care aligned with implementation science frameworks ^23–27^ and factors (barriers and facilitators) influencing their implementation ^28–32^, China remains in a very early, foundational phase. Thus, the aims of this study were: (1) to identify the barriers and facilitators experienced by Chinese healthcare professionals in implementing core OA treatments; and (2) to understand the current clinical practices in OA management within the Chinese clinical context.

## PATIENTS AND METHODS

### Study Design

This study employed a mixed-methods design to investigate the barriers and facilitators experienced by Chinese healthcare professionals in implementing core OA treatments and to understand their current clinical practices in OA management. Mixed-methods research is particularly suited to these aims, as it provides both depth and breadth of understanding^33^. An exploratory sequential approach ^34^ was selected, considering the limited literature on OA care delivery in the Chinese clinical context and the approach’s comparatively more robust validity ^35^. This design also allowed for the iterative exploration of key themes, and the validation of findings, across a broader population. This study proceeded in three phases: initial semi-structured interviews, a qualitative survey with open-ended questions, and a final quantitative survey evaluation. This study follows the GRAMM (Good Reporting of A Mixed Methods) Study Checklist ^36^, and ethics approval was obtained from the local ethics committee. All participants provided informed consent.

### Participants

Participants in this study were selected based on the healthcare professionals targeted by Chinese clinical guidelines for OA. To ensure representation of diverse specialties contributing to OA care, healthcare professionals managing OA across mainland China were recruited without limitations on specific disciplines or institutions. Demographic variables of participants were collected for all phases, included the type of medical institution (public or private), professional specialty, years of professional experience, OA-related certifications or qualifications, and other relevant information. The detailed selection process for participants in each phase is summarized below.

### Qualitative phase 1

This phase aimed to initially explore the barriers and facilitators to implementing exercise therapy for patients with OA.

#### Participants

Participants at this phase were recruited through professional networks between September 2023 and November 2023. Inclusion criteria required participants to have a minimum of two years of clinical experience in OA-related care and without conflict of interests with the research group. We purposively recruited healthcare professionals with diverse roles, geographical locations, and institutional affiliations to ensure variation in clinical experience and practice settings. There were no further exclusion criteria.

#### Data collection

The interviews were guided by a framework based on the TDF (Theoretical Domains Framework), which offers a structured approach to examining behavioral determinants in healthcare^37^. Interview questions were designed to elicit in-depth responses on participants’ experiences with implementing evidence-based OA practices, barriers encountered, and strategies perceived as facilitators in their clinical settings. The interview guide was included in the **Supplementary material**. Individual interviews were conducted by a trained qualitative researcher (ZW), either in person or via online meetings depending on availability. Interviews were recorded and transcribed verbatim by ZW. Field notes were taken during and after the interviews to capture contextual details. Each interview lasted approximately 40 minutes.

#### Data analysis

We analyzed the data using framework analysis^38^, facilitating the systematic organization of data into themes that reflected participants’ clinical experiences and perspectives. Two researchers (XC and SD) independently coded the transcripts using NVivo software, followed by a collaborative meeting to refine the coding scheme and resolve discrepancies. ZW mapped the finalized themes deductively to the domains of the TDF. Recruitment continued until data saturation was achieved, defined as the point at which no new themes were identified in subsequent interviews.

### Qualitative phase 2

This phase aimed to validate and complement the findings from individual interviews, and to inform the design of further quantitative survey.

#### Participants

Participants for this phase were recruited through flyers with QR code on social media from November 2023 to January 2024. The sample size for this phase was determined using the principle of data saturation, whereby recruitment ceased when no new themes or patterns emerged from the open-ended responses.

#### Data Collection

An online qualitative survey with open-ended questions was conducted to expand the sample size and capture perspectives from a broader group of practitioners across various disciplines in China. In addition to demographic questions, participants were asked to respond to several key questions: (1) “What factors hinder your delivery of exercise-based therapies for your OA patients?”; (2) “What factors facilitate your delivery of exercise-based therapies for your OA patients?” These were followed by a non-mandatory question: (3) “If you do not agree with exercise-based therapies, please provide the reasons behind your decision.” Lastly, participants were asked: (4) “What are your usual treatment options for your OA patients?” and (5) “What are your main treatment principles/goals when selecting treatments for your OA patients?” Participants were required to provide their answers in written text.

#### Data analysis

Two researchers (XC and SD) independently conducted the analysis using summative and deductive content analysis to address both thematic frequency and theoretical depth, which are suitable for analyzing large amounts of open-ended survey data ^39, 40^. Summative content analysis identified and quantified key terms and patterns related to barriers, facilitators, and treatment options for OA. Deductive content analysis then categorized responses into predefined themes derived from the research questions and theoretical frameworks. NVivo software was used to manage and organize data, and discrepancies were resolved through discussion. A senior researcher (ZW) reviewed the final themes to ensure validity and reliability.

### Quantitative phase

This phase aimed to evaluate the applicability of the themes and common practices identified in the qualitative phase by quantifying the most frequently mentioned themes and the most common OA practices in clinical settings.

#### Participants

Participants in this phase were recruited using a combination of convenience and snowball sampling from February 2024 to May 2024. The sample size was determined based on descriptive results reported in similar international study^29^ which reported barriers related to knowledge and skills with proportions ranging from 37% to 88%. At a 95% confidence level, a minimum sample size of 141 was calculated for adequate precision. To allow for subgroup analyses and greater accuracy, a more conservative estimate with a maximum variability of 50% and a 5% margin of error resulted in a required sample size of up to 384 participants.

#### Data collection

A structured survey was developed based on the themes identified in the qualitative phases and piloted with 20 healthcare professionals who previously participated in a local in-person workshop. The final survey (see **Supplementary material**) was administered via the ‘Wenjuanxing’ online platform. The survey primarily included the following measures:

1. Bespoke questions about core OA treatment usage: Respondents answered four questions using a 5-point Likert scale to indicate the frequency of implementing exercise, education, weight management, and support for self-health management. The response options were: “Never”, “Occasionally”, “Half of the time”, “Often”, and “Always”. Higher scores for each question indicate a higher frequency of implementation, with scores calculated separately for each question.
2. Factors impacting the implementation of best practices: Respondents evaluated the applicability of factors impacting best practice implementation using a matrix question with four response options: “Not applicable,” “Somewhat applicable,” “Applicable,” and “Highly applicable.” These factors included time allocation, team-based care, resource availability, guideline accessibility, and patient-related factors, among others.
3. Chinese version of the Osteoarthritis Knowledge Scale (OAKS): This scale assesses respondents’ knowledge of OA management. It consists of 11 items, each rated on a 5-point Likert scale: “False”, “Possibly False”, “Unsure”, “Possibly True”, and “True”. The total score ranges from 11 to 55, with higher scores indicating greater knowledge about osteoarthritis.
4. Items evaluating self-reported confidence in OA care: Based on the OA core capability framework ^41^, respondents rated their confidence in 13 specific capabilities using a 5-point Likert scale: “Not confident at all” (1 point), “Not very confident” (2 points), “Somewhat confident” (3 points), “Confident” (4 points), and “Very confident” (5 points).

#### Data analysis

Data were analyzed using RStudio (version 4.2.0). Descriptive statistics were applied to summarize demographic characteristics, frequencies, and proportions of the quantitative responses. All data were anonymized, with informed consent provided on the opening page, and securely stored, and accessible only to the research team.

## RESULTS

The detailed demographic characteristics, including their professional backgrounds of the participants for all three phases are summarized in **Table 1**.

**Table 1.**
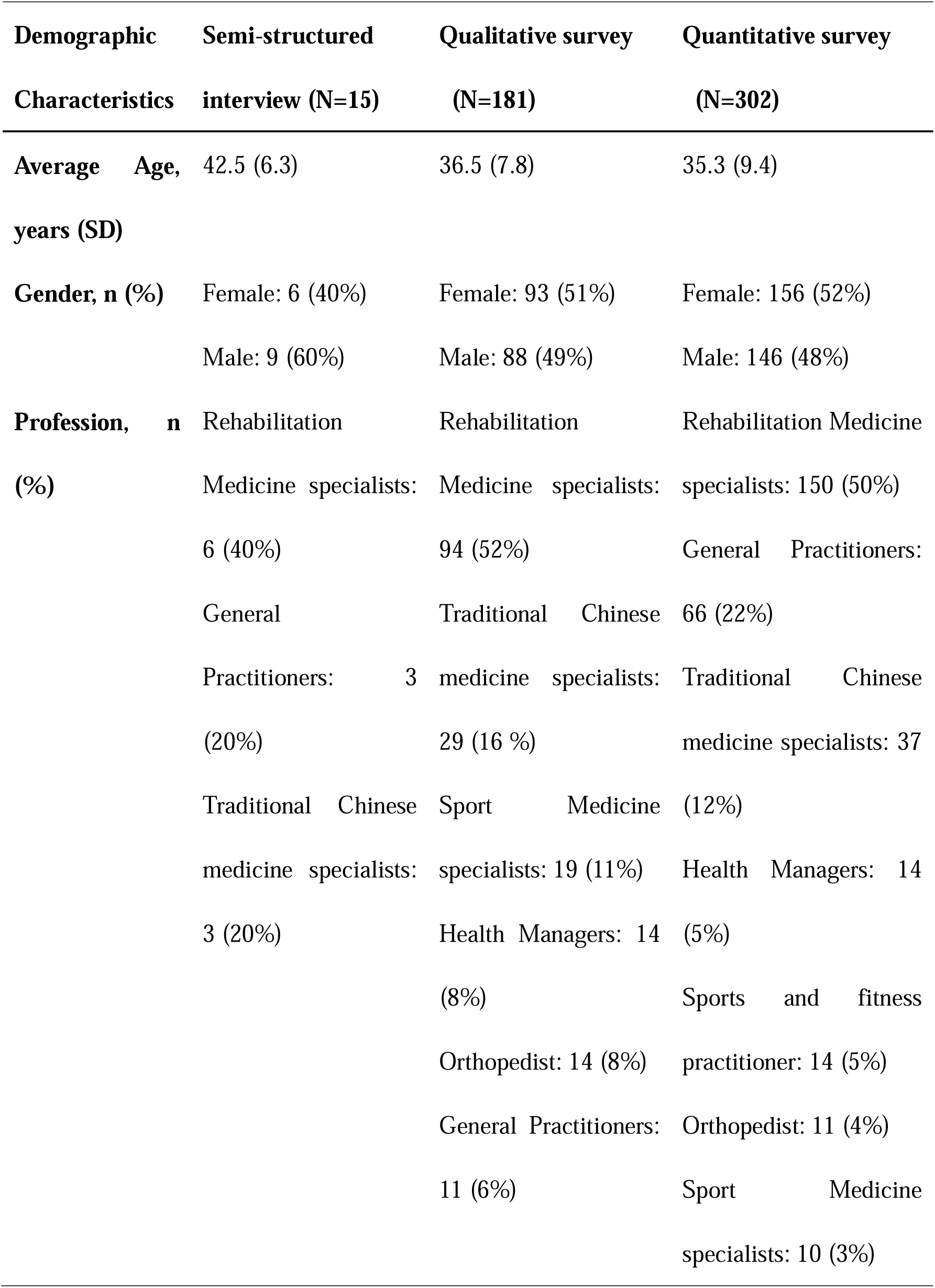
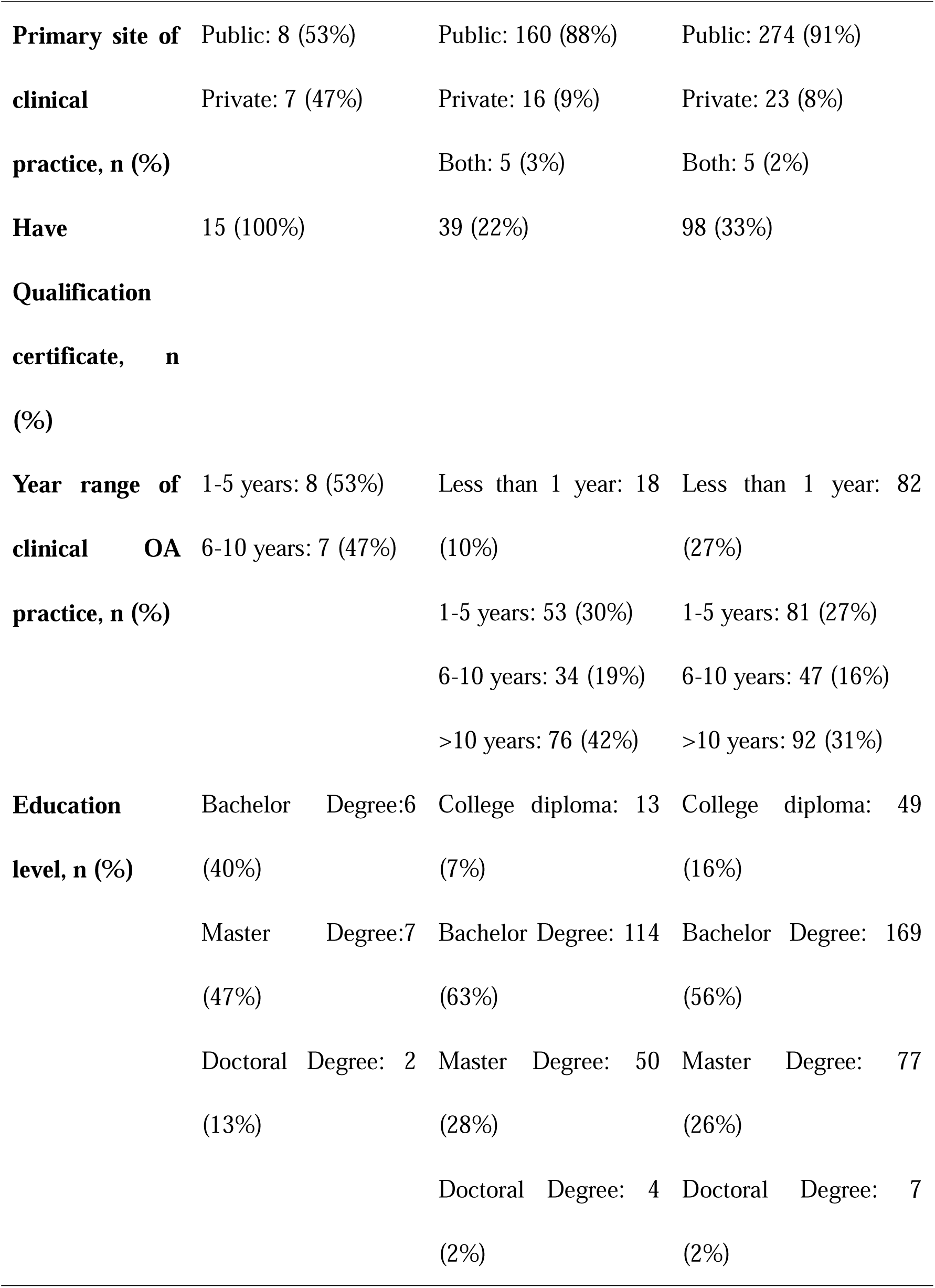
Characteristics of participants.

### Qualitative phase 1

In this initial phase, four themes were identified as barriers and five as facilitators using framework analysis, with the findings presented according to the major TDF domains.

#### Barriers

##### (i) Knowledge gaps and limited awareness of updated evidence

Participants frequently reported challenges in accessing and integrating evidence-based guidelines. Many described a reliance on traditional practices or personal experiences due to a lack of formal training and regular updates. This issue was particularly pronounced in resource-limited settings.

> *“Guidelines are helpful, but in real practice, I rely more on my own experience and what patients respond to.”*

##### (ii) Resource constraints and systemic limitations

Insufficient time, funding, and infrastructure were recurring themes that limited the delivery of evidence-based care. Participants highlighted issues such as low reimbursement rates for exercise therapies and a lack of dedicated rehabilitation facilities.

> *“Our rehabilitation staff are insufficient, facilities and equipment are lacking, communication time with patients is limited, and everyone is exhausted”*

> *“Rehabilitation requires investment in time and equipment, but financial constraints mean we often have to rely on quick fixes, like medication or injections, that are reimbursed”*

##### (iii) Skepticism and resistance to change among colleagues

Resistance to exercise-based interventions, particularly among surgical and pharmaceutical-focused health professionals, was identified as a significant barrier. This skepticism often stemmed from a preference for quicker or more invasive solutions.

> *“Convincing other doctors of the value of exercise is tough; surgery is often seen as the ultimate solution and patients referred to our department are mostly at the end-stage”“Challenging traditional concepts takes time, and inherent habits create obstacles during implementation—for both doctors and the general public”*

##### (iv) Patient misconceptions and adherence challenges

Misunderstanding the role of exercise in OA management was a significant patient-level barrier. Participants noted that many patients associated exercise with potential joint damage or pain, leading to poor adherence to prescribed therapies.

> *“Patients often stop exercising as soon as they feel discomfort, thinking they’re doing more harm than good.” “Discomfort during exercise is often misunderstood; people believe that lying still and avoiding movement is better because pain means something is wrong”*

#### Facilitators

##### (i) Continuous education and professional training

Structured training programs and peer mentoring were recognized as essential for building confidence and competence in implementing evidence-based practices. *“Attending workshops and learning from colleagues has helped me incorporate evidence-based strategies into my daily practice.“*

##### (ii) Technology and digital tools

The integration of telehealth platforms, mobile applications, and exercise videos was highlighted as a valuable facilitator, particularly in under-resourced settings. These tools enhanced patient education and adherence by providing accessible and visual guidance to improve patient understanding of exercise programs.

> *“Apps and demo-videos make it much easier for me to explain exercises and track patients progress remotely and efficiently.” “When I use diagrams and videos, patients seem more willing to follow.”*

##### (iii) Institutional and policy support

Support from healthcare institutions, such as prioritizing rehabilitation services and providing financial incentives for non-surgical interventions, was seen as a crucial facilitator.

> *“When our institution supports exercise therapy, especially when leadership decision-making groups recognize the value of rehabilitation, it makes our job easier.”*

##### (iv) Collaborative teamwork

Multidisciplinary collaboration was frequently cited as a facilitator for implementing OA care. Teams that included physicians, therapists, and support staff were more likely to achieve consistent and effective care delivery.

> *“Working as a team ensures we can align our goals and provide better care to patients.”*

#### Mapping findings to TDF domains

The identified barriers and facilitators were organized into themes based on the relevant TDF domains to ensure a structured analysis. **Table 2** presents examples of barriers and facilitators mapped to each domain.

**Table 2.**
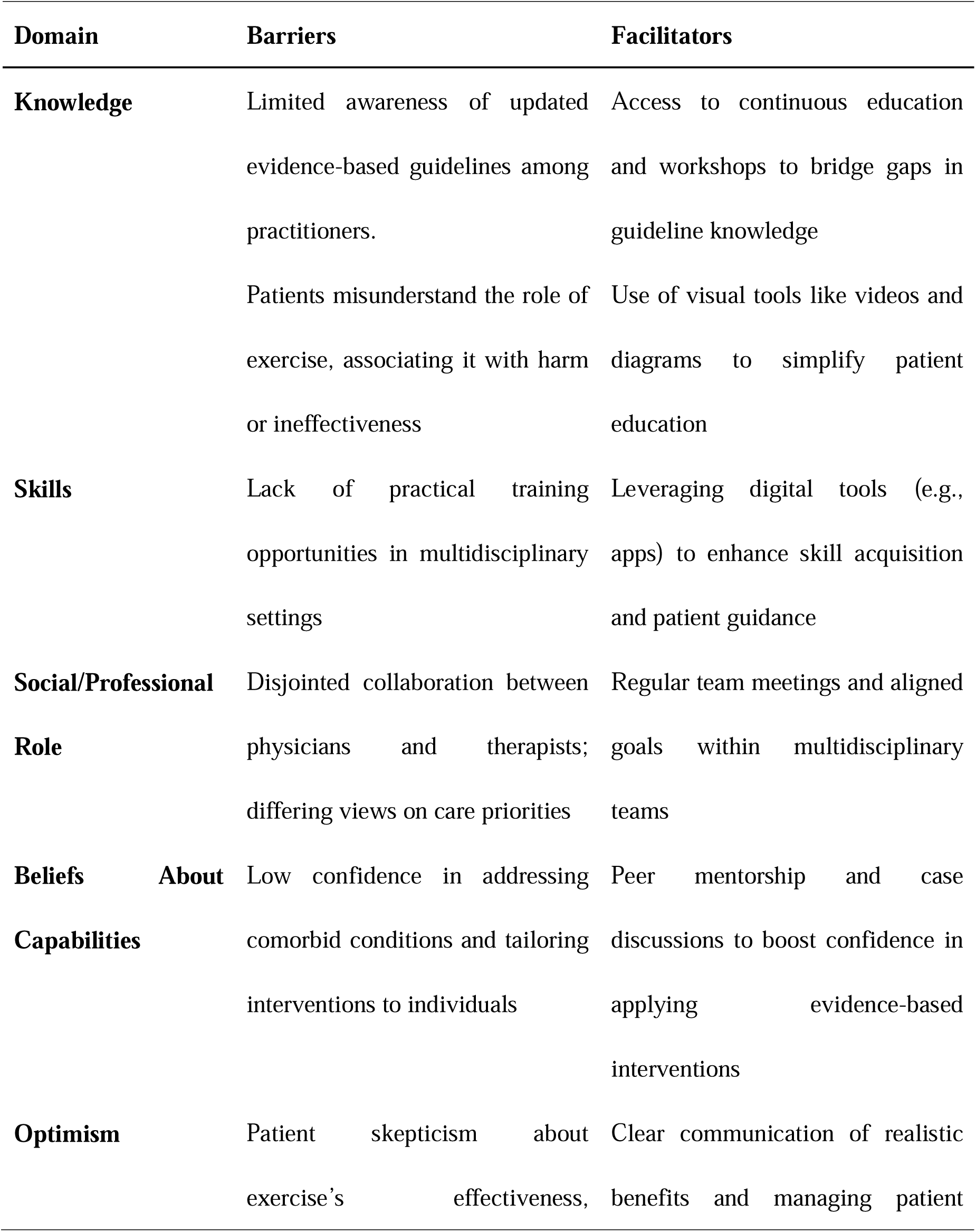
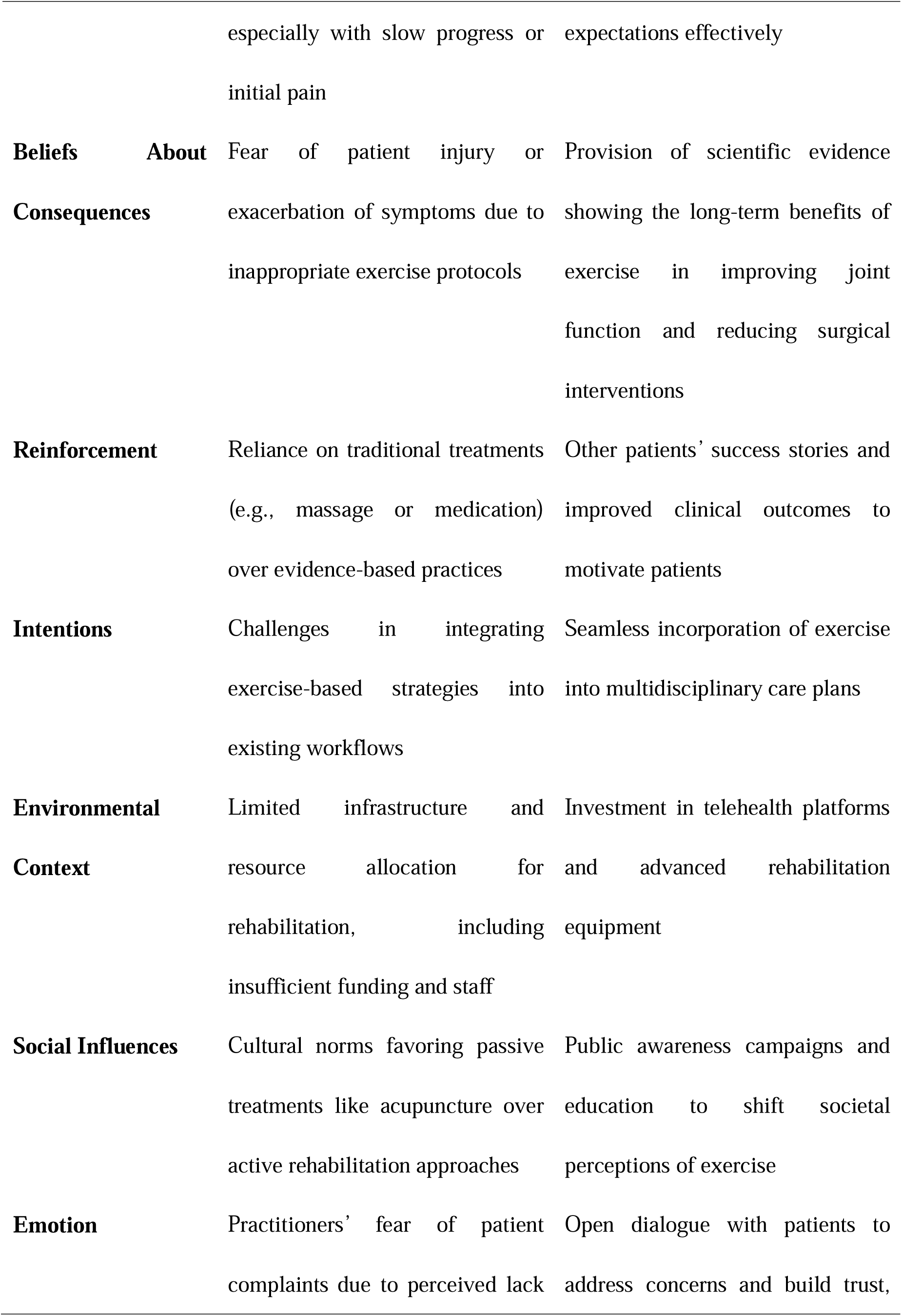
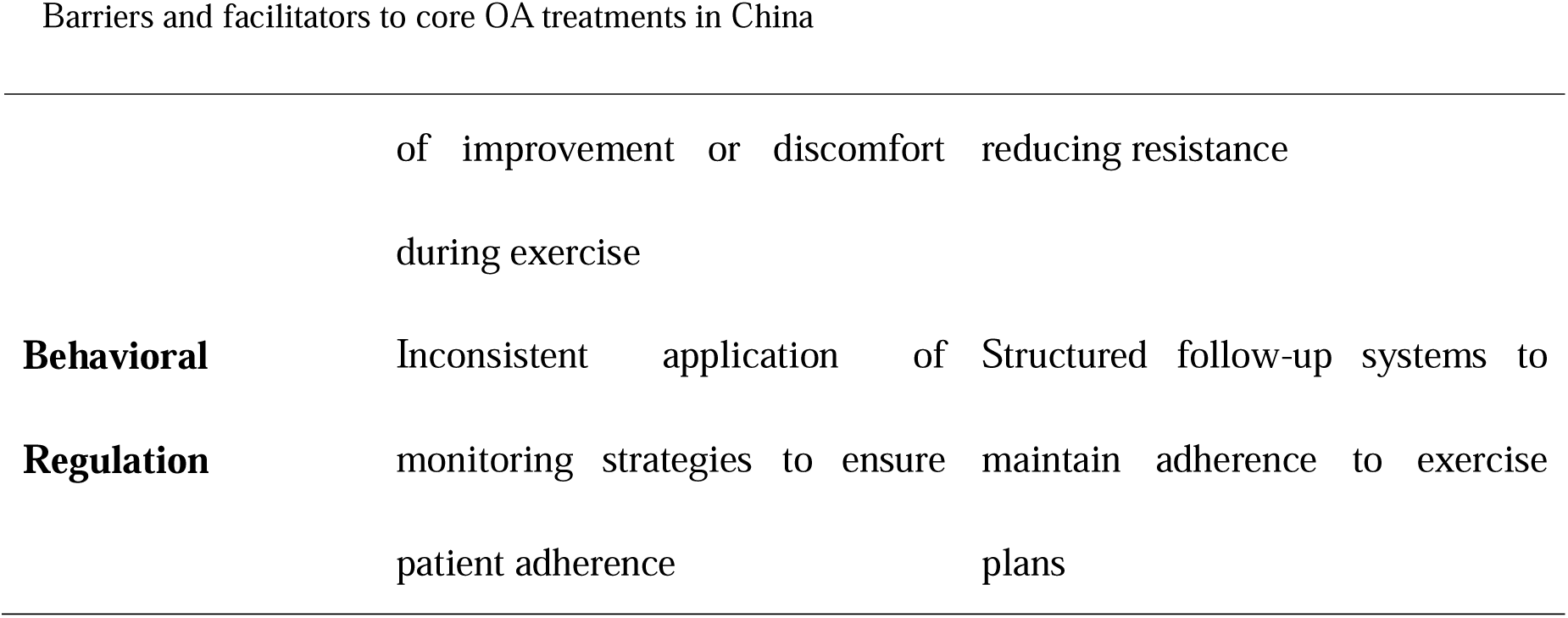
Examples of barriers and facilitators mapped to the TDF from semi-structured interview responses.

### Qualitative phase 2

#### Emerged barriers and facilitators

The qualitative survey (n=181) largely validated the barriers and facilitators identified in Phase 1, while also providing additional insights. For instance, within the ‘Beliefs About Consequences’ domain, participants highlighted that patients’ unrealistic high expectations regarding the speed of symptom relief often led to frustration and early discontinuation of exercise. This finding extended the earlier observations about misconceptions related to exercise. For the ‘Environmental Context’ domain, participants identified a lack of localized easily-understood educational resources tailored to rural populations with low education level, which exacerbated existing disparities in access to evidence-based care. Within the ‘Social/Professional Role’ domain, participants emphasized the importance of clearer task/responsibility allocation and more defined roles within multidisciplinary teams as a facilitator. Additionally, within the ‘Reinforcement’ domain, respondents highlighted the value of incorporating real-world examples of successful delivery to motivate and inspire both patients and healthcare professionals.

Synthesizing the findings from Phase 1 and Phase 2, five themes emerged as key barriers based on their frequency and relevance. These included widespread misconceptions about OA among patients, healthcare professionals, and society at large; limitations in medical insurance policies for non-pharmaceutical services alongside broader economic constraints; insufficient multidisciplinary collaboration with unclear task allocation; a lack of workplace support; and low patient adherence to self-management strategies. Similarly, three themes were identified as facilitators. These included the potential of telehealth and community-based delivery models, the availability of professional training and patient education resources, and the implementation of personalized services supported by positive delivery feedback.

#### Themes of OA treatment goals and usual care practices

Analysis of the text-based responses identified five frequently mentioned themes representing the goals of OA treatment. These themes include pain management, aimed at alleviating patient discomfort; functional recovery and muscle enhancement to improve joint mobility and strength; repair and nourishment techniques focused on cartilage and joint health; anti-inflammatory and swelling reduction strategies; and maintaining overall health through behavioral and lifestyle interventions.

Usual care for OA was also categorized into five main treatment approaches based on the responses. These included physical modality therapies, such as electrotherapy, ultrasound, and similar modalities; TCM practices, including acupuncture and herbal treatments; exercise therapies, emphasizing joint mobilization, functional training, and strengthening exercises; pharmacological treatments, such as Nonsteroidal Anti-Inflammatory Drugs (NSAIDs) and glucosamine; and injection therapies, including corticosteroids and Platelet-Rich Plasma (PRP).

### Quantitative findings

#### Practice of OA management

Participants from 19 provinces in China completed the quantitative survey, with Fujian province accounting for half of the total respondents. As shown in **Table 1**, only 33% of participants completing the survey held a relevant professional healthcare qualification. Among these, 70% held a specialist certification, while the remaining 30% held a physician’s license. The most commonly used OA treatments were physical agent therapy (56%) and TCM (22%), while exercise therapy was utilized in only 9% of cases. Pharmaceutical treatments and injections accounted for 8% and 4%, respectively.

The most frequently reported frequency of managing OA patients in clinical practice was reported as “often/at least once per week”. More than half of the participants reported a delivery frequency of no more than “half of the time” for exercise/physical activity (73.2%), patient education (65.5%), and self-management support (68.2%). The average OA knowledge score was 31.2 (SD = 8.9) out of 55, with 96% of the participants believing that joints wear down with daily use (mean score = 2.1, out of 5), while the role of scans in diagnosing OA was rated the lowest (mean score = 2.0). Among the 13 domains of the OARSI core capability framework, self-rated confidence was lowest for “referrals and interdisciplinary collaboration” (mean score= 3.4, “somewhat confident”), with none of the domains reaching the level of 4 (“confident”).

#### Most common factors impacting implementation

Guided by themes and codes from the qualitative results, we identified the most relevant factors that participants believe impact the implementation of core OA care in their practice, using a 4-point Likert scale (ranging from “not at all applicable” to “highly applicable”). **Table 3** presents the average rating of each factor, sorted in descending order from high to low.

**Table 3.**
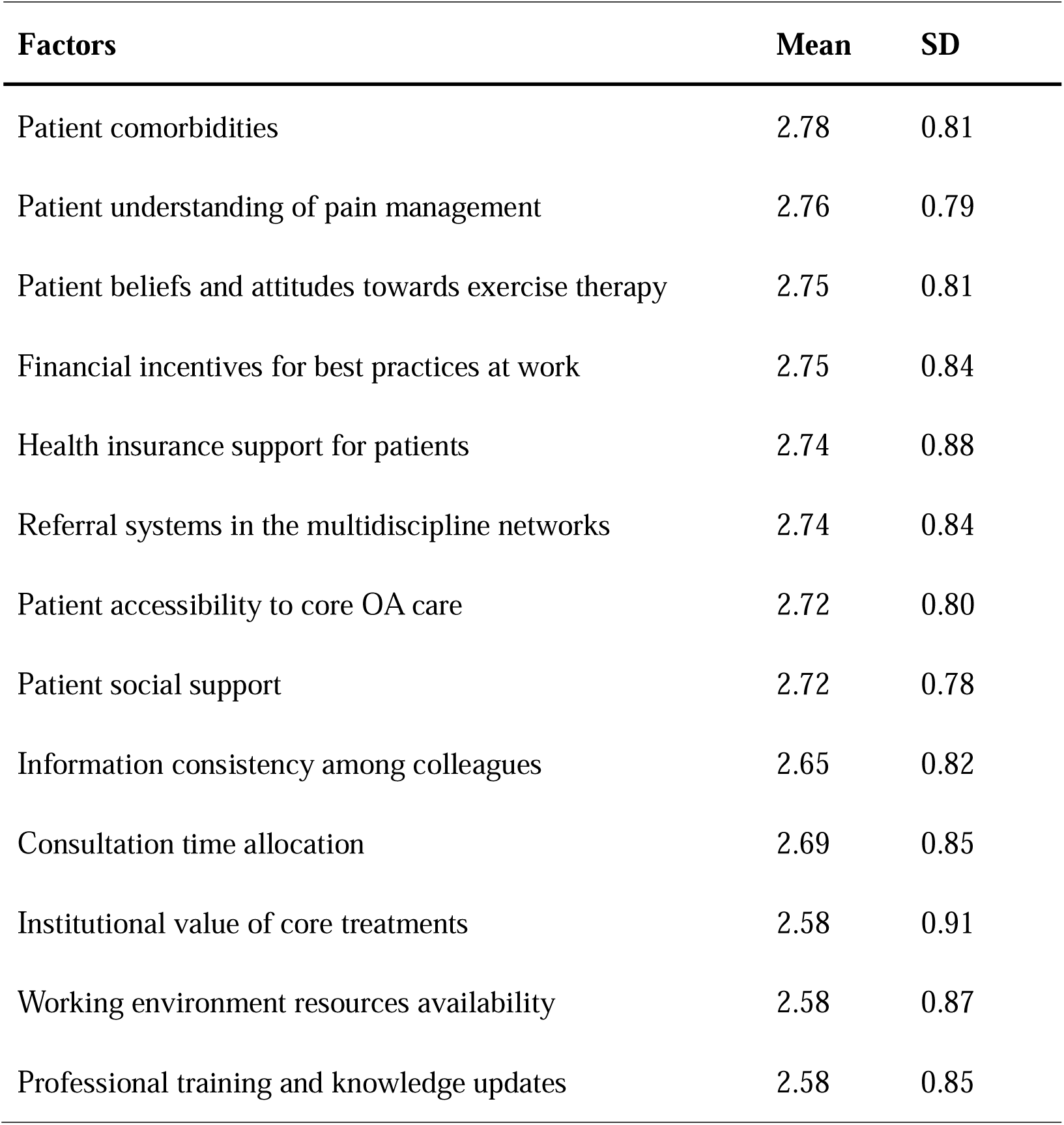
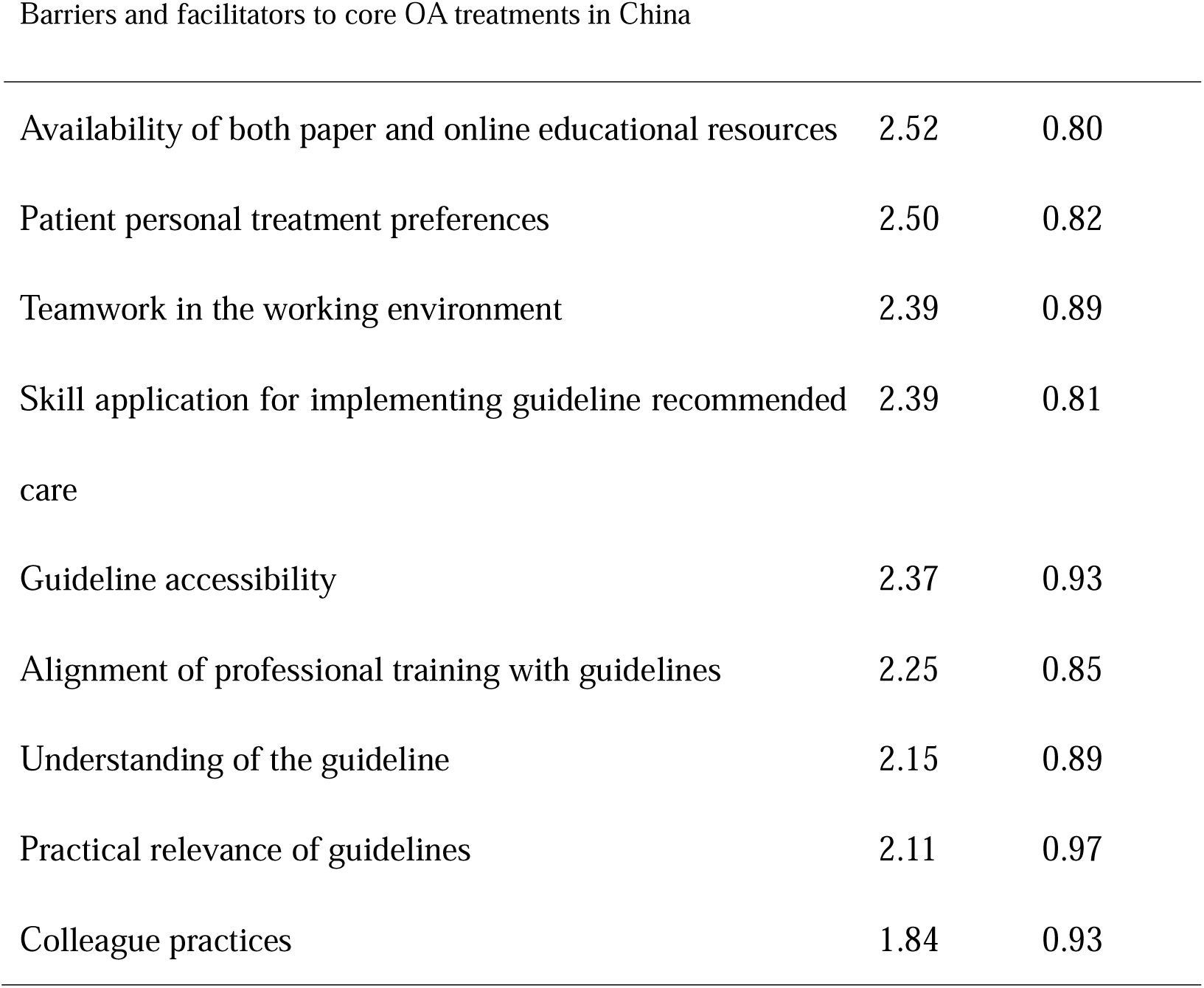
Mean scores of factors influencing the implementation of guideline-recommended core OA treatments.

| **Factors** | **Mean** | **SD** |
| --- | --- | --- |
| Patient comorbidities | 2.78 | 0.81 |
| Patient understanding of pain management | 2.76 | 0.79 |
| Patient beliefs and attitudes towards exercise therapy | 2.75 | 0.81 |
| Financial incentives for best practices at work | 2.75 | 0.84 |
| Health insurance support for patients | 2.74 | 0.88 |
| Referral systems in the multidiscipline networks | 2.74 | 0.84 |
| Patient accessibility to core OA care | 2.72 | 0.80 |
| Patient social support | 2.72 | 0.78 |
| Information consistency among colleagues | 2.65 | 0.82 |
| Consultation time allocation | 2.69 | 0.85 |
| Institutional value of core treatments | 2.58 | 0.91 |
| Working environment resources availability | 2.58 | 0.87 |
| Professional training and knowledge updates | 2.58 | 0.85 |
| Availability of both paper and online educational resources | 2.52 | 0.80 |
| Patient personal treatment preferences | 2.50 | 0.82 |
| Teamwork in the working environment | 2.39 | 0.89 |
| Skill application for implementing guideline recommended care | 2.39 | 0.81 |
| Guideline accessibility | 2.37 | 0.93 |
| Alignment of professional training with guidelines | 2.25 | 0.85 |
| Understanding of the guideline | 2.15 | 0.89 |
| Practical relevance of guidelines | 2.11 | 0.97 |
| Colleague practices | 1.84 | 0.93 |

Many factors were rated highly including: patient comorbidities (such as obesity, mental health problems, and other chronic diseases) (2.8±0.8); patient knowledge of pain science (2.8±0.8) and exercise therapy (2.8±0.8); financial support for both practitioners (2.8±0.8) and patients (2.7±0.9); multidiscipline networks and referral systems (2.7); patient access to appropriate care and support (2.7); and the time available for each consultation (2.7). Personal bias towards guidelines (1.9) and practices of other working colleagues (1.8) were rated as less applicable.

## DISCUSSION

This study is the first to identify Chinese healthcare professionals’ views on the barriers and facilitators to implementing core OA treatments (exercise, patient education, and weight management) through a mixed-methods design. It also explored usual OA clinical practices in Chinese healthcare settings and professionals’ confidence levels in relevant core capabilities. The findings provide a solid basis for informing further interventions to optimize high-value OA health service delivery in China.

### Gaps in current OA care practices

Similar to findings from other countries worldwide, OA management in China remains suboptimal, with few healthcare professionals actively delivering first-line core treatments. Instead, physical modality therapy and TCM were the most commonly used approaches among the participants in our study. A report published in 2013 ^42^ highlighted that physical therapy in China was dominated by these two treatments at that time suggesting little change in OA clinical practice over this time. A similar trend of little change over a decade was observed among Australian general medical practitioners (GPs)^43^. Our findings also revealed that most participants, including many rehabilitation specialists, lacked confidence in their ability to deliver recommended OA treatments. In contrast, a multinational (Australia, New Zealand, Canada) interprofessional study^44^ found that all disciplines, except nurses, reported moderate levels of confidence. This discrepancy may highlight a gap in confidence among our participants. A validated OA Knowledge questionnaire revealed a low level of knowledge in our sample of health care professionals in China, showing certain bias and misconceptions towards OA concepts and managements that have also been observed in other countries ^45^. Given existing evidence-practice gaps in current OA practices in China, it is important to develop strategies to optimize the timely use of core treatments, by addressing the barriers and facilitators identified.

### Implications of identified barriers and facilitators

The barriers and facilitators identified in this study are mainly consistent with those observed in other countries, which typically fall into three meta-themes: personal factors (including both patients and healthcare professionals, knowledge/skills), workforce environment (resources and training), and system-level factors (society and policy) ^28, 29, 46^. For example, the most frequently mentioned and common barrier identified in this study was the widespread misconceptions about OA and exercise therapy among society, patients, and healthcare professionals, which is a common issue worldwide. Therefore, China should develop knowledge dissemination initiatives, such as a toolkit ^47^ with essential components and practical resources for health care professionals and patients.

Environment-level barriers identified in this study are shaped by China’s unique healthcare context. First, restrictions in medical insurance policies for non-pharmaceutical services have led to insufficient funding support for rehabilitation within the healthcare system. The identified issue of multidisciplinary collaboration by our participants is also tied to the limited influence of rehabilitation departments in public hospitals, making referral systems and the role of first assessors/first contact practitioners focused on conservative treatments less effective. This aligns somewhat with a study ^48^ in Australia, which found that the likelihood of general practitioners referring patients to physiotherapists is low for certain health conditions. As the first assessor often plays a central role in managing OA ^49^, optimizing referrals to rehabilitation-led care could be an important way to promote better patient health outcomes. In public hospitals, exercise-based services bring minimal financial incentives, thus rehabilitation departments focus more on TCM than on exercise interventions, which makes organizational leadership support for exercise-based therapies essential.

Facilitators identified in this study highlight the potential importance of tele-health and CHCs. As CHCs have gradually become major primary care providers in China, and OA is a complex chronic health condition requiring long-term self-management support, developing a community-based model of OA care in the future could help alleviate outpatient workload stress in public hospitals^50^. Regarding the use of telehealth in OA care, recent evidence^51^ has shown that telerehabilitation with a physiotherapist can be as effective as in-person care for the non-surgical management of OA. Improving access to core treatments is particularly important, as there is a persistent shortage of rehabilitation specialists in China, and the uneven distribution of rehabilitation resources remains a significant challenge ^20^. Most rehabilitation resources are concentrated in large cities and hospitals, while rural and underdeveloped areas lack sufficient rehabilitation services, even though symptomatic knee OA is more common in rural China ^52^. Telehealth therefore has the potential to bridge this gap and make core treatments more accessible to underserved populations. To facilitate its implementation, relevant training programs should be continuously provided for Chinese to upskill their telehealth knowledge and practices.

This study has several limitations. First, due to the network recruitment process, participants in the quantitative phase were primarily rehabilitation doctors and therapists from Fujian province, which may introduce certain perspective biases, as surgeons or orthopedic doctors might hold different views. Additionally, Fujian, particularly its capital city Xiamen, is a sports-friendly city with strong government support for exercise, which could further influence responses. However, in the preceding qualitative phase, participants were more evenly distributed. Second, different healthcare work environments may present distinct barriers, potentially influencing participants’ beliefs. Since we did not collect detailed information about their specific workplaces, we cannot ensure that the participants’ work settings were evenly represented. Third, this study focused primarily on the management of OA, without addressing other chronic diseases or cases with multiple comorbidities. Finally, we did not collect cost-related economic data in this study, as most participants lacked access to their institutions’ health insurance systems. This aspect will be targeted in future interventional studies. Core OA treatments are infrequently implemented in China, reflecting significant evidence-practice gaps. Targeted efforts are needed to strengthen interdisciplinary collaboration, develop adaptive Chinese OA educational resources for all stakeholders, and provide systematic workplace support for practitioners. Improving accessibility through innovative approaches such as telehealth and community-based care is particularly critical for addressing disparities in resource-limited areas. Future interventions addressing these gaps will optimize evidence-based OA care delivery and improve patient outcomes in China.

## Data Availability

All data produced in the present study are available upon reasonable request to the authors

## Acknowledgments

We sincerely thank Prof. Kim Bennell and Prof. Rana Hinman from the University of Melbourne for their valuable suggestions and feedback during the preparation of this manuscript.

This study was approved by the Ethics Committee of the First Affiliated Hospital of Xiamen University (XMFHIIT-2023SL140), and was funded by the National Social Science Fund of China [23BTY117]. All authors contributed to drafting the manuscript or revising it critically for important intellectual content and approved the final version for submission with no conflict of interests.

ChatGPT was used solely for spelling and grammatical checks of the manuscript.

