## Supplementary material for "Barriers and facilitators to implementing core osteoarthritis treatments in China: a mixed-method study"

**Interview Guide at Phase 1**

| **Category** | **Questions** |
| --- | --- |
| **Knowledge** | - How do you typically describe osteoarthritis to your patients during practice? How do you explain the benefits of exercise to them?  - What is your perspective on evidence-based medicine and clinical guidelines?  - How do you view the recommendations for first-line therapies compared to adjunctive therapies?  - Are you aware of the advantages and disadvantages of different treatment methods? |
| **Skills** | - What skills do you think are necessary to better provide exercise interventions?  - Are there any tools that you find particularly helpful? |
| **Social/Professional Role and Identity** | - Do you think there is goal alignment within your department or specialty regarding osteoarthritis care? |
| **Beliefs About Capabilities** | - Compared to others, how do you evaluate your ability to deliver exercise interventions?  - Do you feel confident in providing targeted plans rather than routine approaches?  - Are there any uncertainties you encounter during the diagnosis and treatment process? |
| **Optimism** | - How optimistic are you about patients achieving improved outcomes through exercise interventions and self-management programs? |
| **Beliefs About Consequences** | - What do you think are the outcomes of using exercise interventions (e.g., do the benefits outweigh potential risks)?  - What outcomes might result from other treatment methods?  - Do you have any concerns about the consequences of using conservative therapies? |
| **Reinforcement** | - What motivates you to use exercise interventions?  - Do you rely on evidence from literature or the recommendations of leaders and colleagues? |
| **Intentions and Goals** | - Does using exercise interventions and self-management support tools (e.g., internet technology, referrals) affect your original treatment goals? |
| **Attention and Decision Processes** | - What factors influence your decision-making process when considering the provision of exercise intervention services?  - How likely are you to offer an exercise intervention plan? |
| **Environmental Context and Resources** | - Are there factors such as healthcare pathways, wages, working hours, or environmental support that impact your ability to provide exercise interventions?  - Do you think internet tools or other supportive resources would help you deliver exercise interventions more effectively? |
| **Social Influences** | - What social factors in China do you think currently impact the decision-making and provision of exercise interventions? |
| **Emotion and Behavior Regulation** | - How would you feel if exercise intervention became a mandatory approach?  - How do you think other doctors would feel?  - Do you have any concerns about this? |

**Quantitative survey**

Title: Survey on the implementation status of core therapy for osteoarthritis

Section 1: Personal Information

1. Your gender (Single choice)

Female

Male

2. Your age (Open-ended)

3. What type of healthcare system are you currently working in? (Single choice)

Public health system

Private healthcare institution (e.g., private clinic, private hospital)

Both

Other

4. Your highest level of education (Single choice)

Below undergraduate

Undergraduate

Master’s degree

Doctorate

Other

5. Your profession (Single choice)

Rehabilitation medicine/physical therapy

Sports medicine

Orthopedics

Traditional Chinese medicine

Rheumatology

Public health practitioner (please specify)

Health manager/Nursing

General practitioners

Sports science practitioner

Other (please specify)

6. Years of experience related to osteoarthritis healthcare (Single choice)

Internship (please specify the duration)

Less than 1 year

1–5 years

6–10 years

More than 10 years

Not related

7. Frequency of treating knee osteoarthritis patients (Single choice)

Very rarely (once in the past 6 months)

Rarely (2–5 times in the past 6 months)

Occasionally (at least once a month)

Often (at least once a week)

Very frequently (5 or more times a week)

8. The primary city in China where you practice clinical care (Open-ended)

9. Do you hold any national qualifications or certifications related to osteoarthritis care? (Single choice)

Yes (please specify)

No

Section 2: Frequency of Best Practice Implementation

1. How often do you prescribe or implement exercise/activity plans for OA patients? (Single choice)

Never

Occasionally

Half of the time (50% of patients)

Often

Always (for all patients)

2. How often do you provide structured patient education programs for OA patients? (Single choice)

Never

Occasionally

Half of the time

Often

Always

3. How often do you provide self-management support for OA patients? (Single choice)

Never

Occasionally

Half of the time

Often

Always

4. How often do you provide weight management advice for overweight or obese OA patients? (Single choice)

Never

Occasionally

Half of the time

Often

Always

5. Do you agree with the following strategies to promote exercise adherence? (0 = strongly disagree; 10 = strongly agree)

Setting goals and plans related to exercise interventions

Explaining the potential benefits of exercise to patients

Discussing barriers to exercise adherence with patients and addressing them

Keeping an exercise journal (paper-based or via apps)

Encouraging the use of reminders (e.g., internet tools or manual reminders by healthcare workers)

**Section 3: Factors Impacting Best Practice Implementation**

(Matrix single choice: Not applicable, Somewhat applicable, Applicable, Highly applicable)

Consultation Time Allocation: The amount of time allocated per consultation for delivering recommended non-drug interventions.

Colleague Practices: The extent to which clinical colleagues adhere to best practices.

Multidisciplinary Care Implementation: The feasibility of implementing team-based or multidisciplinary clinical management in the workplace.

Working Environment Resources Availability: Availability of infrastructure, equipment, information, administrative support, and staff for supporting best practice implementation.

Access to Training and Knowledge Updates: Opportunities for continuous training and access to resources for updating clinical knowledge and skills.

Financial Incentives for Best Practices: Availability of financial incentives to support the implementation of best practices.

Institutional Value of Core Treatments: The perceived value of exercise therapy and self-management support within the institution.

Insurance Support: Availability of financial or insurance support to facilitate treatment decision making.

Referral Network: Access to structured healthcare networks for appropriate and effective referrals.

Accessibility of Core Treatments in the System: Availability of core evidence-based therapies compared to other treatment options.

Information Consistency Among Colleagues: Coordination and consistency of information provided by healthcare workers.

Guideline Awareness: Familiarity with core OA therapies recommended by clinical guidelines.

Skill Application for Core Treatments: The ability to apply specific skills to deliver recommended therapies effectively.

Alignment of Training with Guidelines: The alignment between prior OA management training and current guideline recommendations.

Availability of Educational Resources: Availability of resources to educate patients on exercise and pain management strategies.

Guideline Accessibility: Ease of accessing high-quality clinical guidelines.

Understanding of the Guideline: The clarity and comprehensibility of clinical guidelines.

Practical Relevance of Guidelines: The extent to which guidelines align with practical clinical scenarios and patient characteristics.

Reliance on Experience and Guidelines: Balancing reliance on personal clinical experience with guideline recommendations.

Patient Treatment Preferences: Patients' preferences for treatments that align with or differ from guidelines.

Patient Resource Availability: Patients’ access to necessary treatments and resources.

Patient Beliefs and Attitudes: Patients’ beliefs and attitudes toward exercise and OA management.

Patient Comorbidities: The influence of patients’ psychosocial health and comorbidities on care.

Patient Understanding of Pain Management: Patients’ knowledge and understanding of pain science.

Patient Perceived Social Obligations and Relevant Support: Patients’ other responsibilities in the their society in comparison to the support they receive

Section 4: Core Competencies for Quality OA Care

1. Self-assessment of confidence in core competencies (based on OARSI framework):

For each competency, please rate your confidence level:Not confident at all; Not very confident; Somewhat confident; Confident; Very confident

Competencies:

Effective communication

Person-centered care

Medical history taking

Physical examination

Assessment and diagnosis

Developing care plans and intervention strategies

Providing prevention and lifestyle advice

Delivering self-management strategies

Understanding therapeutic modalities

Knowledge of surgical interventions’ pros and cons

Interdisciplinary collaboration and referrals

Evidence-based practice and quality improvement
